# Interpreting discordant SARS-CoV-2 diagnostic test results

**DOI:** 10.1101/2023.02.07.23285547

**Authors:** Oluwaseun F. Egbelowo, Spencer J. Fox, Graham C. Gibson, Lauren Ancel Meyers

## Abstract

We introduce a model to interpret discordant SARS-CoV-2 test results and estimate that an individual receiving a positive rapid antigen test followed by a negative Nucleic Acid Amplification Test had only a 12-24% chance of being infected in the United States from March 2020 to May 2022.

## Text

Throughout the COVID-19 pandemic nucleic acid amplification tests (NAATs) and rapid antigen tests (RAT) have been widely used to direct patient care and control transmission (1). While NAATs, like RT-PCR, tend to have higher sensitivity and specificity than RATs (2), they are often more costly and take significantly longer to process than RATs (3,4). As of the end of 2022, 22 RATs are available in the United States. They are increasingly used across the US for at-home symptom-based testing and asymptomatic screening in healthcare, educational, and public event settings (5).

Between June 2020 and April 2022, healthcare providers often recommended confirmatory NAAT testing following a positive RAT, given the high false positive rate for RAT’s when community disease prevalence is low (6,7). When a patient receives a negative NAAT result following a positive RAT result, clinicians must decide which of the two results is likely erroneous and suggest a course of action. In this study, we present a statistical model to guide the interpretation of discordant test results that considers test sensitivity and specificity as well as the estimated prevalence of the virus in the community (Appendix).

As a case study, we consider SARS-CoV-2 tests that were widely used in 2021, with estimated RAT sensitivity of 84.6% and NAAT false negative rates of 68%, 37%, 24%, and 21% depending on whether the NAAT is administered 0, 1, 2, and 3 days after the RAT test, respectively (2). For a patient that receives a positive RAT followed by a negative NAAT test, we estimate the probability that the RAT is erroneous and the patient is, in fact, not infected (Figure 1A). This probability is greater than 80% if community prevalence is below 200 new weekly COVID-19 cases per 100,000 people, which is the CDC threshold for low community prevalence (8) and generally declines as disease prevalence increases (Figure 1A). There is a trade-off between NAAT accuracy and speed of diagnosis. If the two tests are given on the same day, then the RAT false positive probability is 80% [95% CI: 44.6%-100%] or 89% [95% CI: 62%-100%] at community prevalences of 0 to 0.3% or 0 to 500 new weekly cases per 100,000, respectively. If, instead, the NAAT is administered three days after the RAT, the corresponding probabilities increase to 86.7% and 96.4%, respectively. Our confidence in the negative NAAT result peaks when the NAAT is administered four days after the RAT (Appendix Figure 2). Barring other external information (e.g. symptomicity), clinicians can be 80% and 64% confident that the initial RAT positive result was a false positive when COVID-19 cases per 100,000 people are 200 and 500 new weekly cases for Low and Medium/High respectively.

**Figure 1.**
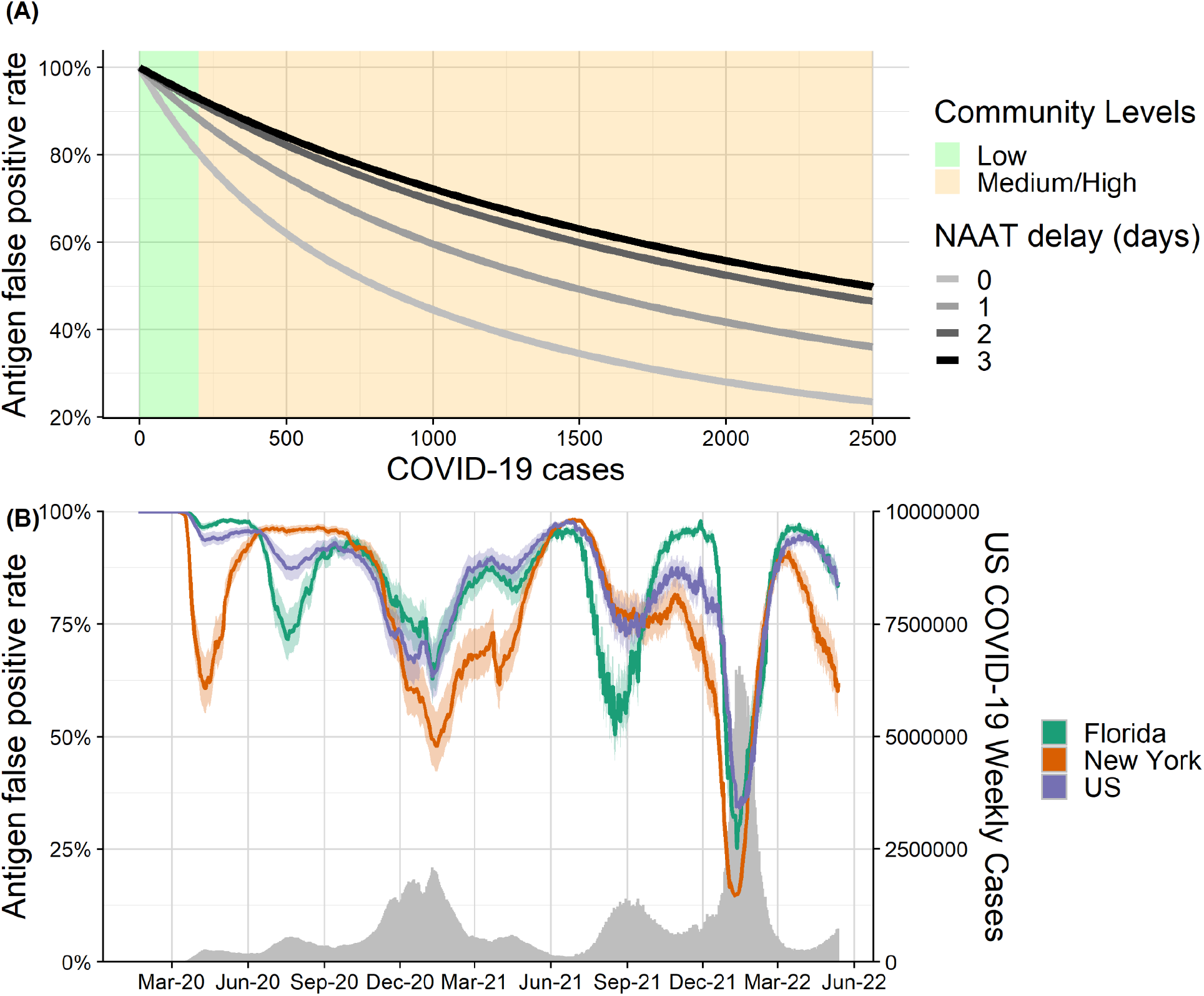
Estimated probability that positive RAT result is erroneous given a subsequent negative NAAT test, for different levels of community transmission. (A) Estimated RAT false positive probability for levels of community transmission ranging from 0 to 500 COVID-19 cases per 100,000 people. The green and orange shading correspond to the CDC’s threshold for Low and Medium/High community levels (8). Line color corresponds to different numbers of days between the initial RAT test and confirmatory NAAT test, ranging from same day (lightest gray) to three days later (black). (B) Estimated RAT false positive probability for the US (purple), Florida (green) and New York (red) from March 2020 to May 2022, assuming the NAAT is administered one day after the RAT. Shading reflects uncertainty in CDC’s estimated COVID-19 infection underreported, ranging from 1 in 3 to 1 in 5. The gray time series along the bottom indicates the daily seven-day sum of reported COVID-19 cases in the US.

Between May 2020 and May 2022, we estimate that RAT false positive probability in the US ranged from 34% to 97% new weekly cases per 100,000 people, assuming a 25% case reporting rate (Figure 1B) (9). For patients receiving conflicting test results (i.e., positive RAT followed by negative NAAT), the probability of an erroneous RAT was lowest during the Omicron surge during the winter of 2021-2022. At the peak, we estimate RAT false positive probabilities of 15% (95% CI 11%-20%), 25% (95% CI 21%-32%), and 34% (95% CI 29%-41%) for New York, Florida, and the US as a whole (Figure 1B).

Rapid and reliable diagnoses of severe infectious diseases is critical for both clinical care and infection control. Yet, the first two years of the COVID-19 pandemic revealed enormous barriers to deploying inexpensive, rapid and accurate tests to combat a newly emerging or rapidly evolving pathogen. We developed this intuitive framework in the fall 2021 to guide decision-making by individuals, physicians and public health officials in the Austin, Texas metropolitan area. It informed the University of Texas’ decision on when a clinician’s visit may be needed. Although our case study assumes test accuracies and case reporting rates specific to the first two years of COVID-19 transmission in the US, the inputs can be readily modified to guide the interpretation of discordant tests as COVID-19 continues to evolve and new pathogen threats emerge (10).

## Data Availability

Data produced in the present work are contained in the manuscript and available online at
https://raw.githubusercontent.com/nytimes/covid-19-data/master/us-counties.csv

## Acknowledgements

We acknowledge the financial support from the National Institutes of Health (grant no. R01 AI151176) and the Centers for Disease Control and Prevention (grant no. U01IP001136).

## About the Author

Dr. Oluwaseun Egbelowo is a postdoctoral researcher in the Department of Integrative Biology at the University of Texas at Austin. He uses the application of mathematical and statistical techniques to aid in decision-making for the control of infectious diseases. Dr. Spencer Fox is an Assistant Professor at the University of Georgia in the Department of Epidemiology & Biostatistics. His research interests include statistical modeling of emerging infectious diseases.

## Appendix Estimating the probability of a false positive RAT, given a positive RAT followed by a negative NAAT

Our goal is to estimate the probability that a positive antigen test was a false positive conditional upon a subsequent negative NAAT result. Using Bayes’ theorem, this is given by

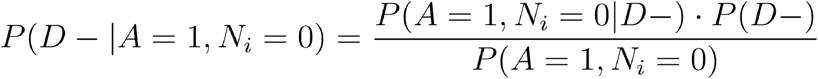

where *D* − denotes the disease state of the individual (minus and plus indicate uninfected and infected, respectively), *A* denotes the result of the antigen test and *N*_*t*_ denotes the result of the NAAT test when administered *t* days after the antigen test (zero and one indicate negative and positive, respectively). We assume that the antigen and NAAT tests are independent of one another.

We then estimate the false positive rate as given by

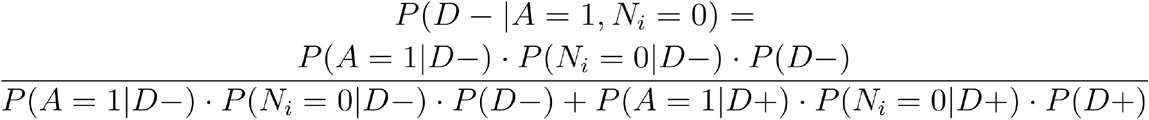

with parameter estimates as described in Appendix Table 1. We assume that is equal to the prevalence of SARS-CoV-2 in the community.

**Appendix Figure 1.**
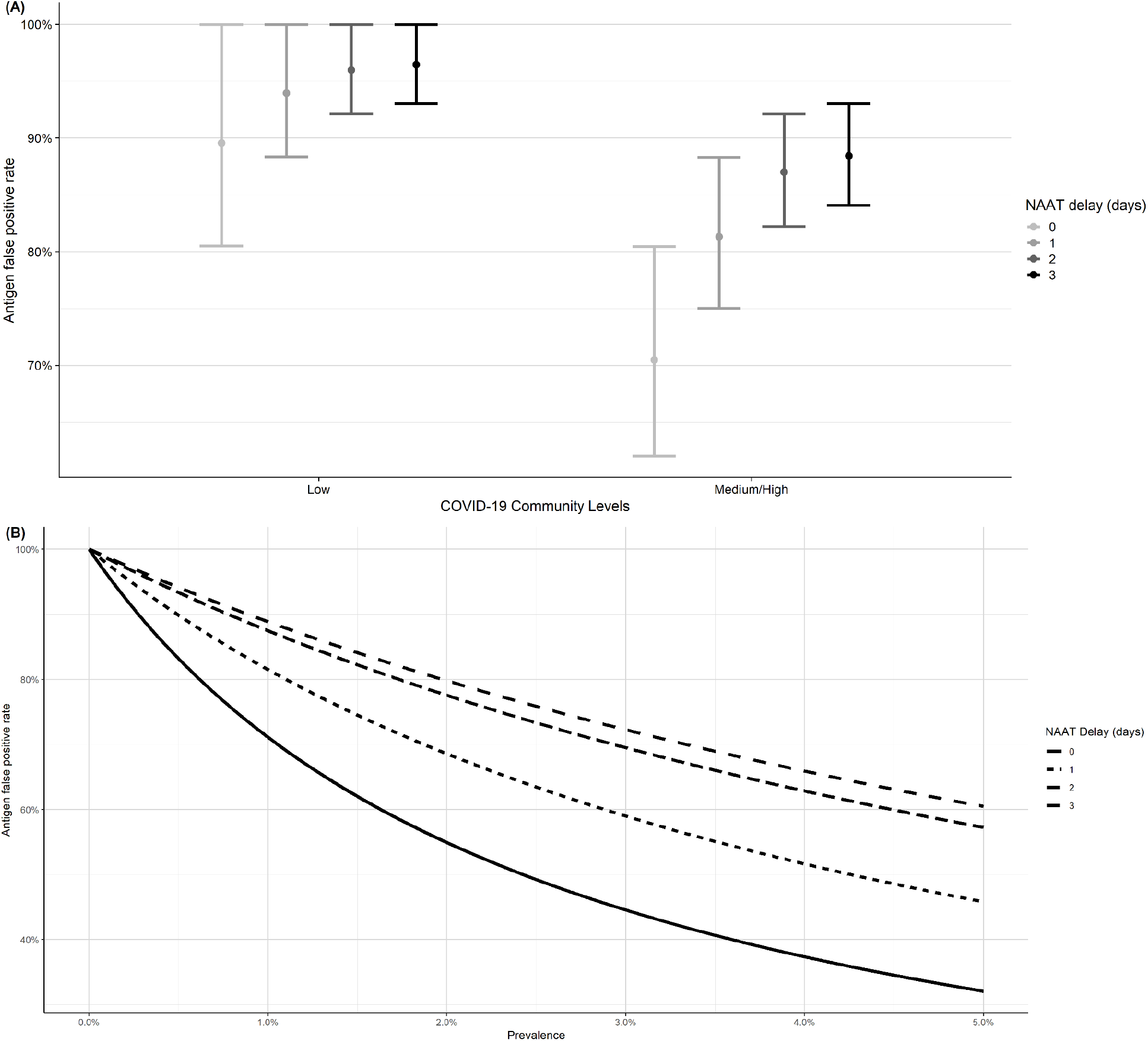
The probability that RAT is a false positive given a subsequent negative NAAT test. (A)The mean probability that less than 200 cases (low) and greater than 200 cases (medium/high) per 100,000 population is RAT false positive given a subsequent negative NAAT test. The error bars are the lowest and highest probability RAT false positive. (B) The probability that a positive RAT is a false positive given a subsequent negative NAAT test, depends on the prevalence of SARS-CoV-2 in the community. Color indicates the number of days between the initial antigen test and confirmatory NAAT.

**Appendix Figure 2.**
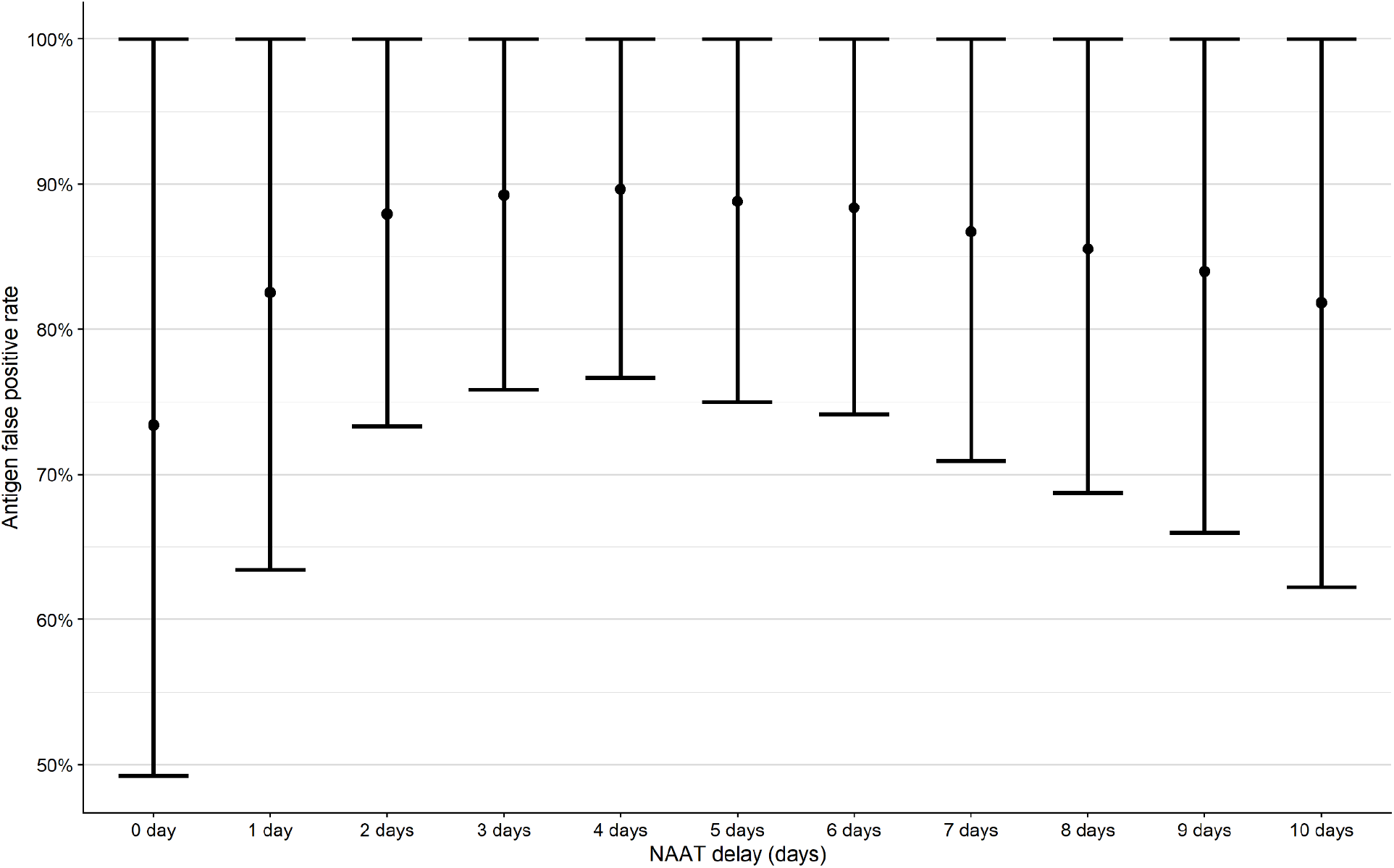
The probability that a positive antigen test is a false positive given a subsequent negative NAAT test with different time delays (3). This shows how sensitive is the delay over time.

**Appendix Table 1.** Model parameters.

| Parameters | Description | Value | Source |
| --- | --- | --- | --- |
| $P(A = 1\vert D-)$ | Antigen test false positive rate | 0.0146 | (1) |
| $P(A = 1\vert D+)$ | Antigen test sensitivity | 0.846 | (1) |
| $P(N_i = 0\vert D-)$ | NAAT true negative rate | 0.98 | (2) |
| $P(N_i = 0\vert D+)$ | NAAT false negative rate | 1 day after antigen test: 0.37 | (3) |
|  |  | 3 days after antigen test: 0.21 |  |
| $P(D+)$ | Prevalence | 0 - 5% | (4) |

**Appendix Table 2:** Thresholds for requiring a clinical visit following a positive antigen test and a negative NAAT confirmatory test. For each confidence level, we recommend requiring doctors’ visits when community admissions exceed the values provided in the table.

| Desired confidence level | Community admissions threshold for requiring clinical confirmation |  |
| --- | --- | --- |
|  | 1 day between antigen and NAAT tests | 3 days between antigen and NAAT tests |
| 50% | 4.3% | 7.6% |
| 65% | 2.4% | 4.2% |
| 80% | 1.1% | 2% |
| 90% | 0.5% | 0.9% |
| 95% | 0.24% | 0.46% |

**Appendix Table 3:**
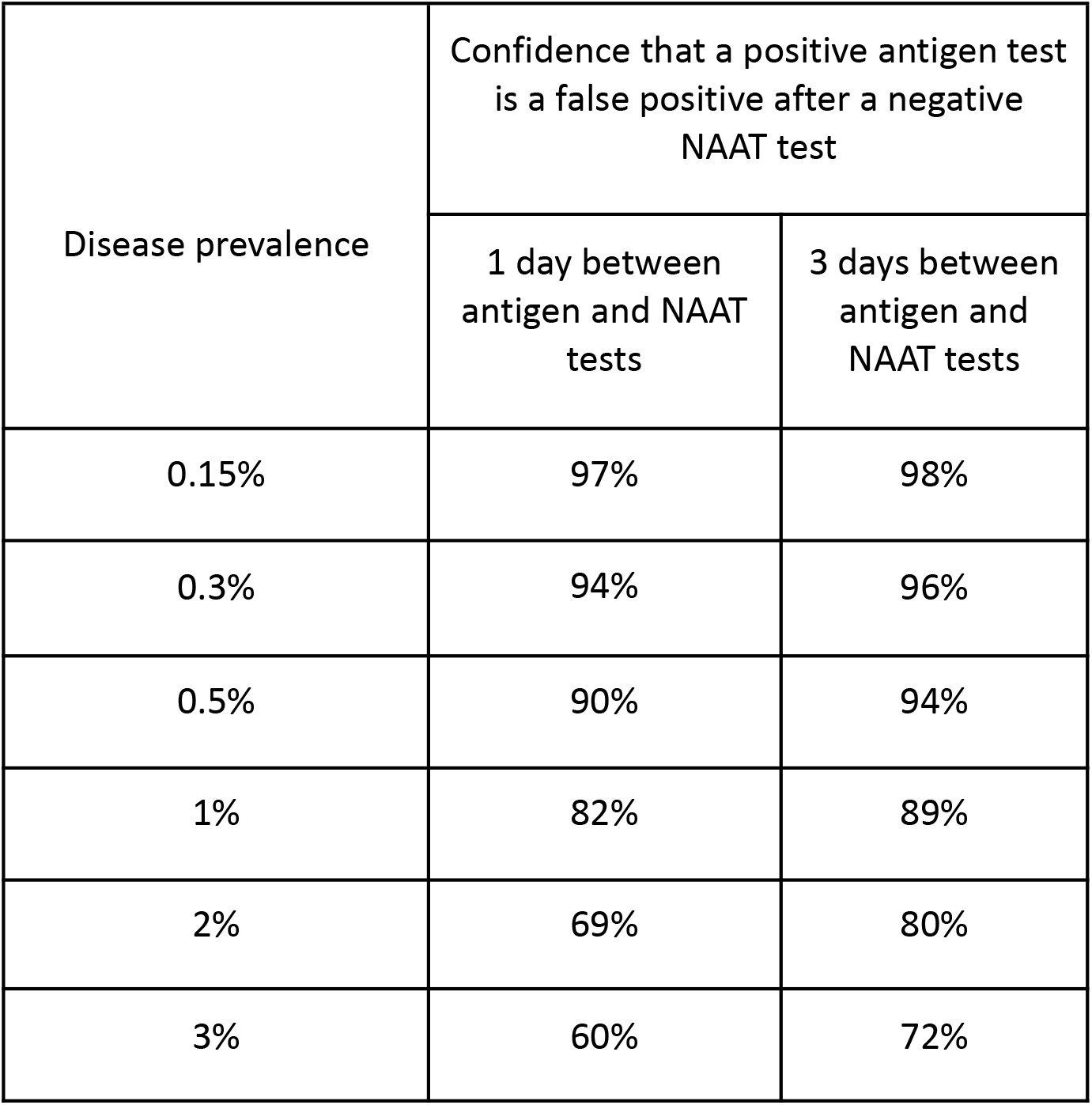
Probability that a positive antigen test is a false positive given a subsequent negative NAAT test, depending on the prevalence of SARS-CoV-2 in the community.

**Appendix Table 4:** Mean probability that a positive antigen test is a false positive given a subsequent negative NAAT test, depending on different community levels.

| Community levels | Mean confidence that a positive antigen test is a false positive after a negative NAAT test |  |  |
| --- | --- | --- | --- |
|  | 0 day between antigen and NAAT tests (95% CI) | 1 day between antigen and NAAT tests (95% CI) | 3 days between antigen and NAAT tests (95% CI) |
| Low | 89.6%<br>(80.5%-100%) | 93.8%<br>(88.1%-100%) | 96.4%<br>(93.0%-100%) |
| Medium/High | 70.5%<br>(62.0%-80.5%) | 80.9%<br>(74.5%-88.0%) | 88.4%<br>(84.1%-93.0%) |

## Notes

### Competing Interest Statement

The authors have declared no competing interest.

### Author Declarations

https://raw.githubusercontent.com/nytimes/covid-19-data/master/us-counties.csv

